# BA.2 omicron differs immunologically from both BA.1 omicron and pre-omicron variants

**DOI:** 10.1101/2022.05.10.22274906

**Authors:** Annika Rössler, Antonia Netzl, Ludwig Knabl, Helena Schäfer, Samuel H. Wilks, David Bante, Barbara Falkensammer, Wegene Borena, Dorothee von Laer, Derek Smith, Janine Kimpel

**Affiliations:** Institute of Virology, Department of Hygiene, Microbiology and Public Health, Medical University of Innsbruck, Peter-Mayr-Str. 4b, 6020 Innsbruck, Austria; University of Cambridge, Center for Pathogen Evolution, Department of Zoology, Cambridge, UK; Tyrolpath Obrist Brunhuber GmbH, Hauptplatz 4, 6511, Zams, Austria

## Abstract

**Background:** Several studies have shown that SARS-CoV-2 BA.1 omicron is an immune escape variant and current vaccines and infection with pre-omicron variants provide limited protection against BA.1. Meanwhile, however, omicron BA.2 has become the dominant variant in many countries and has replaced BA.1. As BA.2 has several mutations especially in the receptor binding and the N terminal domain compared to BA.1, we analyzed whether BA.2 shows further immune escape relative to BA.1.

**Methods:** We characterized neutralization profiles against the new BA.2 omicron variant in plasma samples from a variety of individuals with different numbers of exposures to infection/vaccination, including samples from previously virus-naïve, BA.2 omicron-infected individuals. To illustrate antigenic differences of the two omicron sub-variants and pre-omicron variants we performed antigenic cartography and generated antibody landscapes.

**Results:** Unvaccinated individuals after a single exposure to BA.2 had limited cross-neutralizing antibodies to pre-omicron variants and to BA.1. Consequently, our antigenic map, which included all Variants of Concern and both BA.1 and BA.2 omicron sub-variants, showed that both omicron sub-variants are distinct to pre-omicron variants, but that the two omicron variant are also antigenically distinct from each other. The antibody landscapes illustrate that cross-neutralizing antibodies against the whole antigenic space, as described in our maps, are generated only after three or more exposures to antigenically close variants but also after two exposures to antigenically distinct variants.

**Conclusions:** Here, we describe the antigenic space inhabited by the relevant SARS-CoV-2 variants, the understanding of which will have important implications for further vaccine strain adaptations.

## Introduction

During the course of the severe acute respiratory syndrome coronavirus-2 (SARS-CoV-2) pandemic, mutations in the viral genome occurred leading to an evolution from ancestral variants to currently circulating Variants of Concern (VoC). Variants that were selected either had improved viral fitness/transmission kinetics such as the alpha (B.1.1.7) variant, immune escape properties such as the beta (B.1.351) variant or a combination of both such as the delta (B.1.617.2) variant. Most recently, the omicron (B.1.1.529) variant has been described as VoC with the two predominant sub-lineages BA.1 and BA.2 and more recently also BA.4 and BA.5 in South Africa and BA.2.12.1 in the US.

We and others previously showed that the BA.1 omicron variant led to the strongest immune escape so far seen in SARS-CoV-2 variants.^1-4^ While the virus strongly escapes neutralizing antibodies induced by infection with pre-omicron variants or two doses of vaccination, T cell responses seem to be more conserved.^5-7^ However, multiple exposures improve neutralizing antibody titers against BA.1 omicron as seen in individuals after booster vaccination or hybrid immunity.^1,2,8,9^

BA.1 and BA.2 omicron variants share common mutations; however, each has also unique mutations. Some of these unique mutations are located in the receptor-binding domain or the N-terminal domain of the spike protein, both important epitopes for binding of neutralizing antibodies. Therefore, BA.1 and BA.2 omicron might have distinct neutralization profiles. The antigenic difference of pre-omicron, BA.1 omicron and BA.2 omicron variants is supported by a study showing distinct profiles of sensitivity against therapeutic monoclonal antibodies.^10^ Initial reports analyzing sera from wild type convalescent or two dose vaccinated individuals show that both omicron subvariants escape neutralizing antibody responses to a similar degree perhaps with a trend of lower escape by BA.2. A booster immunization or hybrid immunity strongly enhances neutralizing antibodies against both omicron variants.^11-13^ However, these studies mainly analyzed samples from vaccinated individuals and little is known about neutralizing antibody profiles induced by BA.2 omicron infection in previously naïve individuals.

Antigenic cartography is a tool to visualize antigenic differences between different virus variants. Several maps have been described showing pre-omicron variants and also including the BA.1 omicron variant.^14-17^ The position of the BA.2 omicron variant has so far been only added to maps using sera from infected hamsters but no human data are available.^18^

In the current study, we characterize neutralization profiles against the new BA.2 omicron variant in plasma samples from a variety of individuals with different numbers of exposures to infection/vaccination, including samples from previously naïve BA.2 infected individuals, and use these data to generate an antigenic map including all current VoC.

## Methods

### Ethics statement

The ethics committee (EC) of the Medical University of Innsbruck has approved the study with EC numbers: 1100/2020, 1111/2020, 1330/2020, 1064/2021, 1093/2021, 1168/2021, 1191/2021, and 1197/2021.

### Patient characteristics

We included individuals with single, double or three and more exposures. Details regarding age, sex, vaccination status etc. of participants are given in the Supplementary Methods and Table S1.

### Neutralization assay

A focus forming neutralization assay for SARS-CoV-2 has been used as previously described.^1,19^ Continuous 50% neutralization titers were calculated in GraphPad Prism 9.0.1 (GraphPad Software, Inc., La Jolla, CA, USA) using a non-linear regression. Titers <1:16 were considered negative. Titers <1 were set to 1 and titers >1:16,384 were set to 1:16,384. The following replication competent SARS-CoV-2 virus isolates have been used: D614G (Isolate B86.2, GISAID ID EPI_ISL_3305837); alpha variant (B.1.1.7, isolate C69.1, GISAID ID EPI_ISL_3277382); alpha variant with E484K mutation (C79.2, GISAID ID EPI_ISL_3277383); beta variant (B.1.351, isolate C24.1, GISAID ID EPI_ISL_1123262); gamma variant (P.1.1, isolate hCoV-19/Germany/BY-MVP-000005870/2021, GISAID ID EPI_ISL_2095177); delta variant (B.1.617.2, isolate SARS-CoV-2-hCoV-19/USA/NY-MSHSPSP-PV29995/2021, GISAID ID EPI_ISL_2290769); BA.1 omicron variant (isolate E16.1, GISAID ID EPI_ISL_6902053); BA.2 omicron variant (isolate E65.1, GISAID ID EPI_ISL_12486408). Viruses were grown on Vero cells stably overexpressing TMPRSS2 and ACE2.

### Antigenic cartography

Antigenic maps were constructed from single infection and double vaccination sera as described in ^14,20^. Using multidimensional scaling, antigen variants and sera are positioned in a lower-dimensional space based on serum antibody titers. For each serum and antigen pair, antigenic distances are calculated from the titer reduction of the antigen against which the specific serum has the highest titer (usually the homologous, infecting antigen) to antigen *i*. For each serum-antigen pair, serum and antigen coordinates are optimized to minimize the error between the Euclidean distance in the map and the target distance from the measured titers, where one antigenic unit in the map corresponds to one two-fold dilution of titers in the neutralization assay. The *Racmacs* package ^21^ was used to create antigenic maps with 1000 optimizations, a dilution stepsize of 0, the minimum column basis set to “none”, and titers below the limit of detection of 16 set to “<16”. The reactivity of the P.1.1 variant was reduced by one two-fold due to as high or higher than homologous titers in all but the BA.1 and BA.2 convalescent serum groups (Figure S1). A two-dimensional map was suitable to represent the antigenic relationships as assessed by map diagnostics (Supplementary Methods).

### Antibody landscapes

First infection and double vaccination antibody landscapes were constructed as described in ^14^ using the P.1.1 reactivity adjusted map as base map. Different to previous approaches,^22^ reactivity of single exposure sera was assumed to adopt a cone-like shape, with its apex at the serum coordinate and its height equal to the maximum titer, decreasing at a constant rate of one two-fold per antigenic unit (slope = 1 log2 unit). The assumption of a slope = 1 does not pertain to more cross-reactive sera exposed to multiple variants, hence the serum coordinates and landscape slope for multi-exposure sera, excluded from the initial map, were fitted. Using R’s ^23^ *optim* function with the parameters maxit=500 and the optimization method “L-BFGS-B”, the error between Euclidean map distance and measured titer distance was minimized to obtain x-and y-coordinates for each serum and the landscape’s slope per serum group. P.1.1 measured titers were reduced by one two-fold to account for likely assay reactivity bias. The GMT landscapes show the average of all individual serum landscapes per serum group. For GMT calculation, reactivity bias of individual sera was accounted for as described in.^14^

### Statistical analysis

Statistical analysis was performed using non-parametric repeated measures ANOVA with Friedman’s test for multiple comparisons.

## Results

First, we analyzed neutralizing antibody profiles in previously naïve individuals after an infection with the BA.2 omicron variant. All of these individuals had detectable neutralizing antibodies against the BA.2 omicron variant itself, however neutralizing antibodies against pre-omicron and BA.1 omicron were only occasionally above the limit of detection (IC_50_ >1:16) and in the few positive samples generally low (Figure 1A). This was in concordance with our previous data where unvaccinated individuals after a pre-omicron VoC infection induced mainly neutralizing antibodies against pre-omicron variants but not BA.1 omicron and vice versa, sera from unvaccinated individuals recovered from BA.1 omicron variant infection mainly neutralized BA.1 omicron but not pre-omicron variants.^1,2^

**Figure 1.**
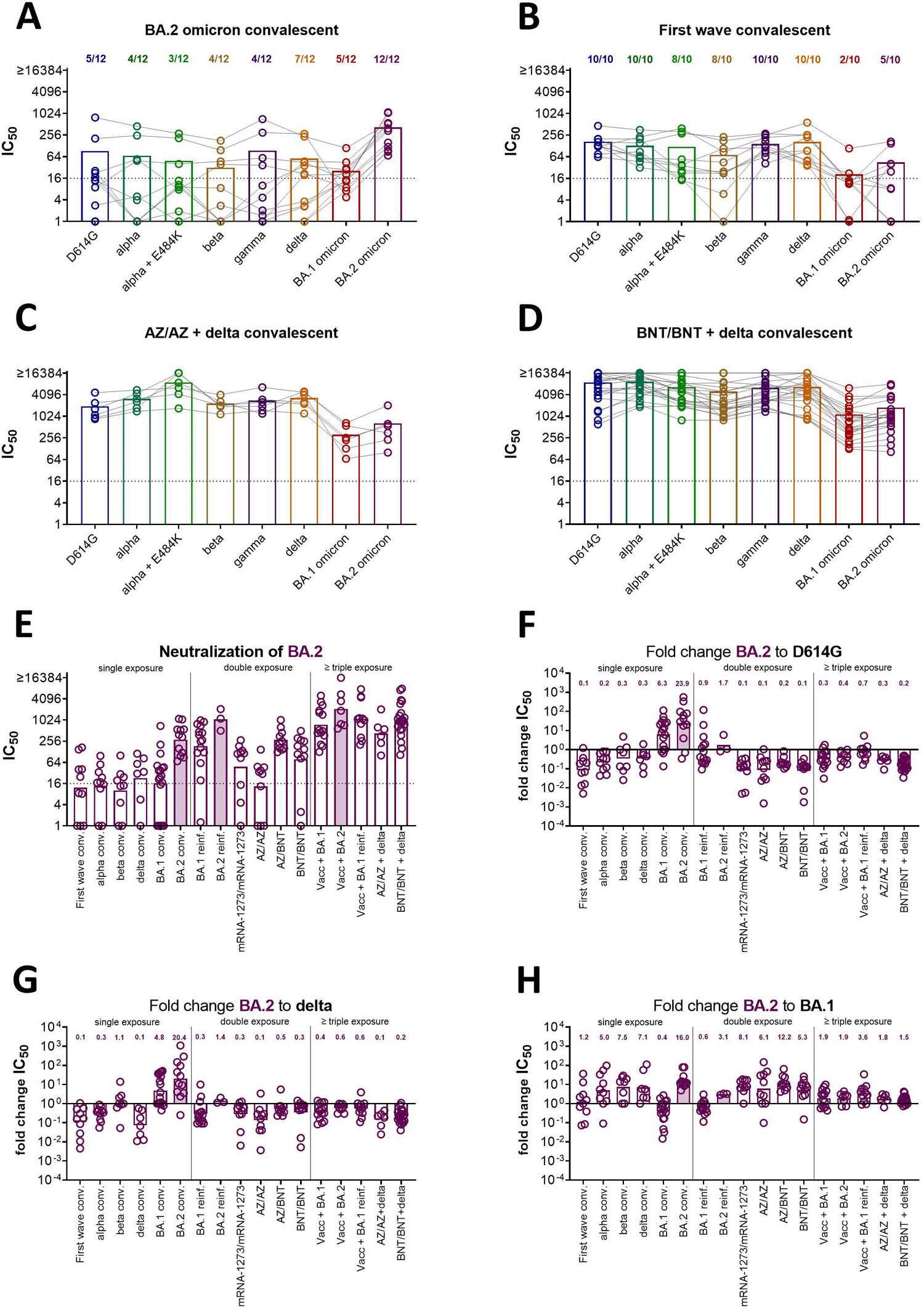
BA.2 omicron has a neutralization profile distinct from both pre-omicron variants and BA.1 omicron. Plasma samples were collected after a BA.2 omicron (n = 12, Panel A) or wild-type infection (n = 10, Panel B) in previously naïve patients (no vaccination and no known history of prior infection) or in patients with hybrid immunity, Panel C two doses of ChAdOx-S1 followed by delta infection (n = 6) and Panel D two doses of BNT162b2 followed by pre-omicron variant (presumably delta) infection (n = 22). Samples were analyzed for 50 % neutralizing antibody titers (IC_50_) against D614G, alpha, alpha + E484K, beta, gamma, delta, BA.1 omicron, and BA.2 omicron. Shown are individual patient samples as circles connected by lines and mean titers as bars. Numbers above bars indicate proportion of positive samples (titers >1:16). Panel E) IC_50_ titers against BA.2 omicron for different groups of individuals with single exposure (non-vaccinated convalescent), double exposure (re-infection with BA.1 or BA.2 omicron after historic non-omicron infection in unvaccinated or two doses of vaccine), and three or more exposures (breakthrough infection after vaccination) were determined. Shown are individual patients (circles) and geometric mean (bars) for each group. Dotted lines in Panel A-E indicate limit of detection. Fold change of IC_50_ against BA.2 relative to D614G (Panel F), delta (Panel G), and BA.1 (Panel H) were calculated. Part of the D614G, delta and BA.1 neutralizing antibody titers used for calculation of fold changes in Panels F-G have been previously published in ^1,2^. Shown are individual patients (circles) and geometric mean fold changes (bars). Numbers above bars indicate geometric mean fold changes.

To complete the dataset, we now also analyzed neutralizing antibody titers against our panel of variants for plasma samples from individuals who had been infected during the first wave in Austria (March/April 2020).^24^ As expected, these individuals had high neutralizing antibody titers against pre-omicron variants with slightly reduced titers against the immune escape variants beta and alpha with E484K mutation (for both variant 8 out of 10 individuals above cut-off). However, neutralizing antibodies against both omicron sub-variants were only induced in part of the individuals (2 out of 10 for BA.1 and 5 out of 10 for BA.2, Figure 1B). In contrast, individuals with hybrid immunity showed a broad neutralizing antibody response against all variants analyzed (Figure 1C, D). This was true for breakthrough infections after two doses of an mRNA vaccination (BNT162b2, BNT) as well as after two doses with a vector vaccine (ChAxOx-1-S, AZ). These breakthrough infections were presumably all caused by the delta variant.

We next analyzed neutralizing antibody titers against the BA.2 omicron variant for a broader selection of samples. We included individuals with single exposure (unvaccinated convalescent from first wave, alpha, beta, delta, BA.1 omicron, or BA.2 omicron variant), with double exposure (unvaccinated after pre-omicron variant and BA.1 omicron re-infection or vaccinated with two doses) or three and more exposures (vaccinated with two or three doses and breakthrough infection). In general, multiple exposures improved neutralizing antibody titers against the BA.2 omicron variant even for individuals that had no contact with the BA.2 omicron variant itself (Figure 1E). Relative to the D614G and the delta variants, neutralizing antibody titers against the BA.2 omicron variant were higher in unvaccinated BA.1 or BA.2 omicron variant convalescent persons but lower in most individuals after pre-omicron variant infection or multiple exposure (Figure 1F, G). However, the drop in neutralizing antibody titers against the BA.2 omicron variant in these groups was not as pronounced as we previously showed it against the BA.1 omicron variant. Additionally, most groups, especially after only one or two exposures, had higher titers of neutralizing antibodies against the BA.2 compared to the BA.1 omicron variant (Figure 1H).

These data indicate that the BA.2 omicron variant is positioned antigenically between pre-omicron variants and the BA.1 omicron variant but distinct to both. To analyze these differences in more detail, we next did antigenic cartography.^20^ In antigenic cartography, antigen variants and sera are positioned relative to each other in a lower-dimensional space based on serum antibody titers, where the distance in the map reflects the dilution steps in the neutralization assay. Applying this approach to the titer data from convalescent and double vaccinated serum groups resulted in a map that could represent the data in 2D well and was robust to assay noise (Figure 2, Figures S2-S10 and Supplementary Methods).

**Figure 2.**
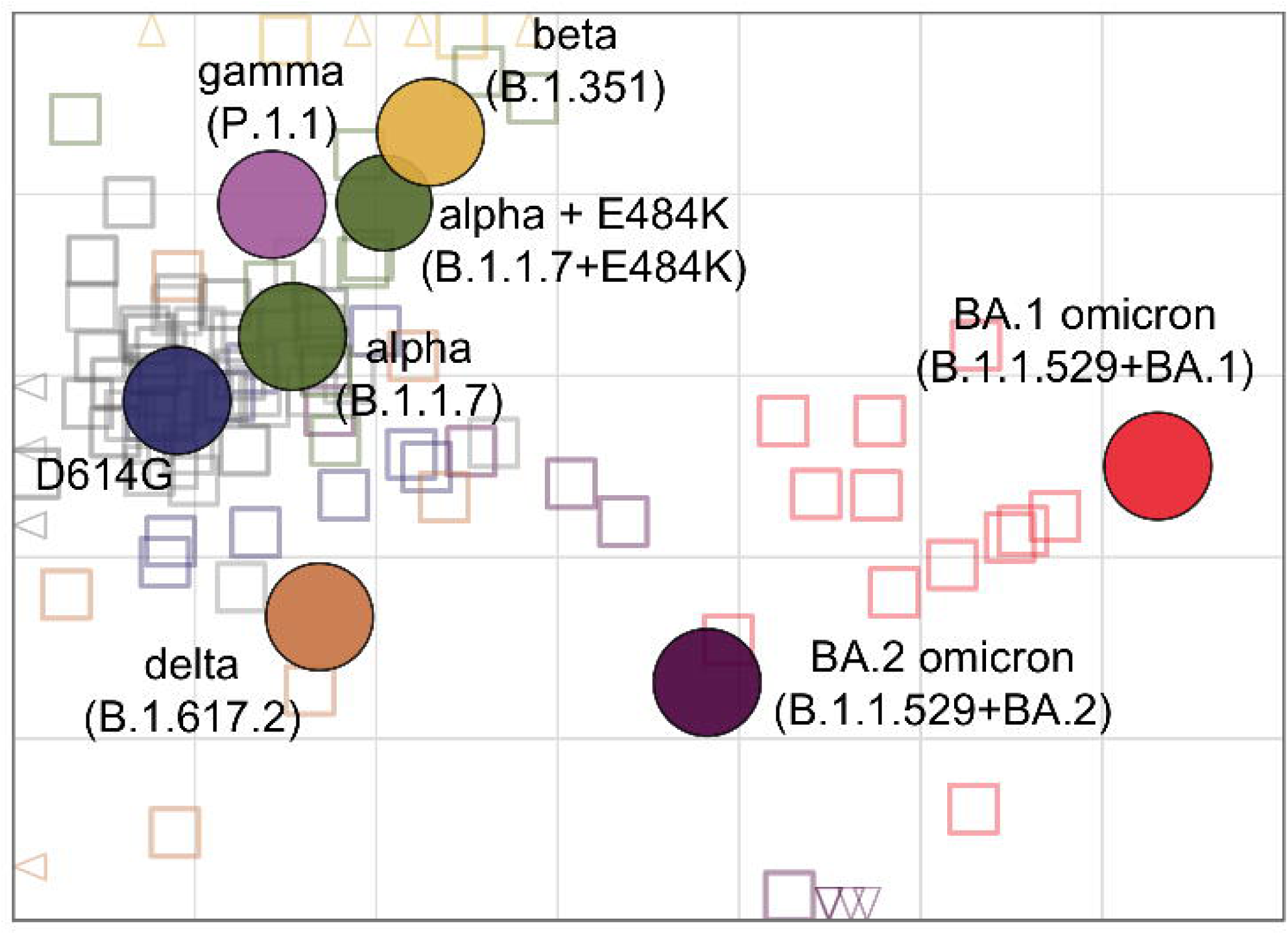
Antigenic map of SARS-CoV-2 variants constructed from single exposure convalescent and double vaccinated sera. Virus variants are shown as colored circles, sera as open squares with the color corresponding to the infecting variant. Vaccine sera are shown in grey tones (from dark to light: mRNA-1273/mRNA-1273, BNT162b2/BNT162b2, ChAdOx-S1/BNT162b2, ChAdOx-S1/ChAdOx-S1). The alpha + E484K variant is shown as smaller circle due to its additional substitution compared to the alpha variant. The x- and y-axis represent antigenic distances with one grid square corresponding to one two-fold serum dilution of the neutralization titer. The map orientation within x- and y-axis is free as only relative distances can be inferred. A non-zoomed in version of the map is shown in Figure S2.

The antigenic map visualizes the substantial difference of the two omicron sub-lineages BA.1 and BA.2 compared to the previously circulating variants (Figure 2) and corresponds well with previous maps.^14,15,18,25^ Wu-1 like variants (D614G, alpha, alpha+E484K), beta and gamma occupy a small space in the map and the delta variant is in roughly the same area. Barely detectable neutralization titers against BA.1 omicron in all but the BA.1 omicron convalescent serum group resulted in its positioning far away from the other variants. As the titer data indicated, BA.2 omicron was located between the pre-omicron variants and BA.1 omicron, approximately equidistant to delta and BA.1 omicron. Low but detectable titers against BA.2 omicron in samples across all serum groups explain the variant’s position. The majority of BA.2 convalescent sera, however, exhibited little cross-reactivity against any other variant (Figure 1A, Figure S11), reflected by their far-off location in the map area (Figure S2).

To visualize the antibody reactivity profile of individuals with distinct infection history, we next constructed antibody landscapes ^14,22^ for each serum group, where a serum’s neutralization titers are plotted above an antigenic map in a third dimension (Figure 3, Figure S11). Grouping by the number of exposures, we found that at least two distinct variant encounters greatly increased reactivity against all other variants. Single variant exposure landscapes had highest reactivity against the infected variant, with similar reactivity profiles of the first wave and delta convalescents and the vaccinated individuals, and alpha and beta convalescents (Figure 3A-B, Figure S11 A-J). BA.1 and BA.2 omicron convalescent landscapes exhibited both unique antibody profiles focused on the area of their root variant, reflecting the observations in the neutralizing antibody titer data.

**Figure 3.**
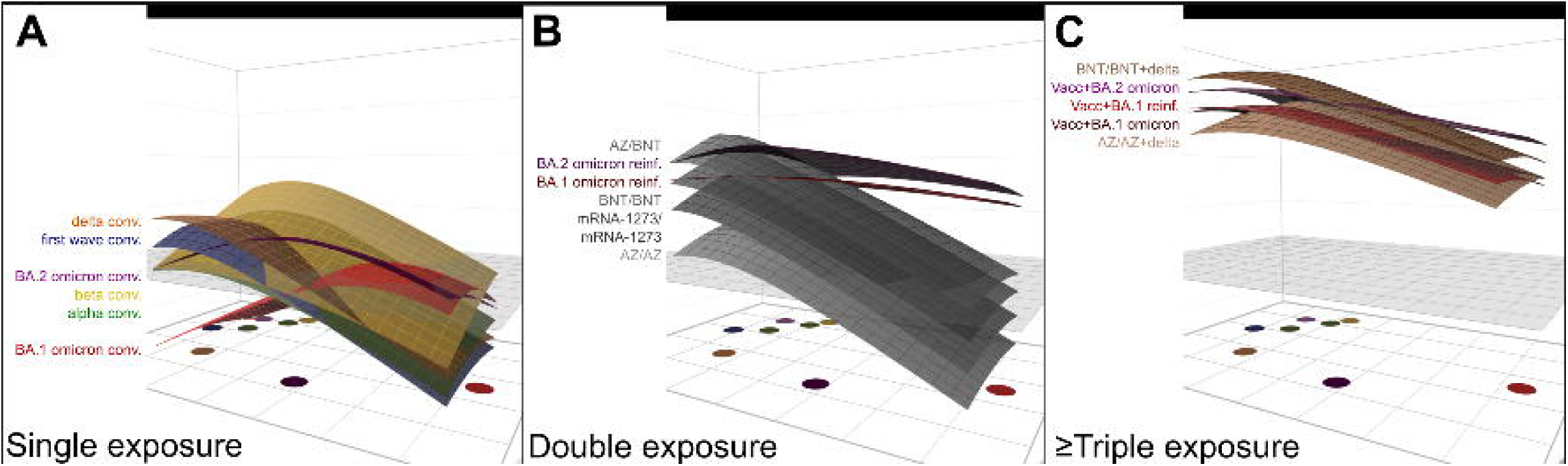
GMT antibody landscapes by number of exposures. The colored surfaces represent the GMT antibody landscapes of the different serum groups. The map shown in Figure 2 serves as base plane, the z-axis reflects log_2_ titers with each two-fold increase marked starting from titer 20. The grey plane at titer 50 serves as reference. Reactivity bias of individual sera was accounted for as described by ^14^. The GMT antibody landscapes were grouped and colored by virus exposures: A Single exposure convalescent sera (from top to bottom : delta (orange), first wave (blue), BA.2 omicron (purple), beta (yellow), alpha (green), and BA.1 omicron (red)). B Double exposure sera (from top to bottom: ChAdOx-S1/BNT162b2 (AZ/BNT), BA.2 omicron reinfection (pre-omicon infection followed by BA.2 omicron reinfection), BA.1 omicron reinfection (pre-omicon infection followed by BA.1 omicron reinfection), BNT162b2/ BNT162b2 (BNT/BNT), mRNA-1273/mRNA-1273, ChAdOx-S1/ChAdOx-S1 (AZ/AZ)). C Triple or more exposure sera (from top to bottom: BNT/BNT + delta breakthrough, Vacc + BA.2 omicron breakthrough, Vacc + BA.1 reinfection (Vacc + pre-omicron breakthrough infection + BA.1 omicron reinfection), Vacc + BA.1 omicron breakthrough, AZ/AZ + deltabreakthrough). Individual sera antibody landscapes are shown in Figure S11.

In contrast, non-omicron infection followed by omicron infection resulted in broad antibody landscapes of similar shape, differing only in magnitude by omicron sub-lineage (Figure 3B). Breakthrough infection with either omicron variant or delta, despite being more similar to pre-omicron variants than to omicron, induced high titers against all variants (Figure 3C, Figure S11 K-N). Further, BA.1 omicron breakthrough and BA.1 omicron breakthrough with previous non-omicron infection landscapes were almost identical. This suggests that, while the increase in cross-reactivity from one to two SARS-CoV-2 variant exposures is substantial, a third variant exposure changes the reactivity profile not to the same extent. Examining this in more detail, we found that the number of exposures increased antibody levels, but the type of exposures impacted the landscape’s shape (Figure S13). On average, antibody reactivity in the multi-exposure cohorts was higher against pre-omicron variants than against the two omicron sub-lineages with the exception of omicron re-infected individuals.

## Discussion

Our results support the hypothesis that the omicron variant is indeed a distinct serotype compared to previous VoC. However, we found the two omicron sub-lineages BA.1 and BA.2 to be antigenically distinct both from each other and from pre-omicron variants resulting in different neutralization profiles and distinct positions on the antigenic map.

To visualize the underlying antigenic relationships of neutralizing antibody data in an antigenic map, it is essential to analyze plasma from double vaccinated individuals or unvaccinated individuals infected with only one variant. Our map is the first to include BA.2 omicron together with BA.1 omicron and pre-omicron variants based on data from human sera using replication competent SARS-CoV-2 variant-based assays. Our findings are in line with a previous study, which used pseudotyped vesicular stomatitis virus and sera from experimentally infected hamsters, and which also found BA.2 omicron to be antigenically distinct from both BA.1 omicron and pre-omicron variants.^18^ We found BA.2 omicron to be closer to the delta variant, whereas their study showed BA.2 omicron to be as distant from delta and beta as BA.1 omicron, however this might be explained by the different host species. Removing BA.2 omicron convalescent sera with especially strong cross-reactivity to other variants, which could potentially be due to undetected prior infection, from our analysis did not change the map (Figure S12). Although previous maps based on human sera lack BA.2, the positioning of the other VOC is highly consistent to the map generated from our data.^14,15^

Looking at how neutralizing antibody reactivity distributed across the mapped antigenic space through antibody landscapes, individuals exposed to a single variant exhibited mainly titers against the specific or closely related variants. A subsequent infection with an antigenically distinct strain did not only increase titers against the new variant, but also against the previous variant and resulted in broad reactivity profiles even to variants the individual was not exposed to. This is in line with other studies where an omicron variant re-infection not only increased neutralizing antibody titers against omicron but also against other variants.^26,27^ Surprisingly, we found that a delta variant breakthrough infection induced high titers against both omicron sub-lineages, although these individuals were never exposed to the omicron variant. In these individuals, neutralizing antibody titers against both omicron sub-variants were comparable to individuals who had a combination of prior infection/vaccination and omicron infection. One limitation of our study is that most samples were collected shortly after infection and therefore we could not analyze longevity of cross-neutralizing antibodies. However, others also found improved levels and durability of cross-neutralizing antibodies after three or more exposures (vaccination or infection).^3,8,27^ In a non-human primate model, titers of cross-neutralizing antibodies were similarly increased by either a third dose of a wild type-specific mRNA vaccine or a beta-specific mRNA vaccine after two doses of a wild type-specific mRNA vaccine.^28^

We propose three hypotheses for the improved cross-neutralization after repeated exposure even for non-exposed variants. Firstly, this could be a consequence of antibody saturation against the encountered virus variants while the reactivity against unencountered viruses could still be boosted with overall increasing antibody titers, resulting in smaller differences in neutralization between exposed and unexposed variants with overall increased antibody titers. Secondly, the prior encounter of two different pre-omicron variants could boost immunity against conserved epitopes shared with the two omicron sub-lineages. Some monoclonal antibodies have been shown to retain activity against BA.1 and/or BA.2 omicron.^29^ Thirdly, the broad polyclonality of the response after exposure to two different variants might contribute to omicron neutralization, as a cocktail of monoclonal antibodies was reported to improve neutralization of BA.1 and BA.2 omicron.^30,31^

Our work presents an important contribution to the discussion on vaccine updates to improve protection against current and future emerging SARS-CoV-2 variants. It suggests, that cross-neutralization improves with repeated exposure and increase in absolute titers of antibodies and that exposure to two distinct variants has protective potential against emerging variants with some degree of similarity to currently and previously circulating VoCs.

## Data Availability

All data produced in the present study are available upon reasonable request to the authors.

## Acknowledgment

We thank Albert Falch, Bianca Neurauter, Eva Hochmuth, Evelyn Peer, Lisa-Maria Raschbichler, Luiza Hoch, Lydia Riepler, and Teresa Harthaller for excellent technical and organizational support. We thank Prof. Florian Krammer and Prof. Viviana Simon for sharing their Delta isolate and Prof. Oliver T. Keppler and Dr. Marcel Stern for sharing their Gamma isolate with us.

JK was supported by the European Union’s Horizon 2020 research and innovation program under grant agreement No. 101016174 and the Austrian Science Fund (FWF) with the project number P35159-B. DS was supported by the NIH NIAID Centers of Excellence for Influenza Research and Response (CEIRR) contract 75N93021C00014 as part of the SAVE program.

## References

1. Rössler A, Riepler L, Bante D, von Laer D, Kimpel J. SARS-CoV-2 Omicron Variant Neutralization in Serum from Vaccinated and Convalescent Persons. N Engl J Med 2022. DOI: 10.1056/NEJMc2119236.

2. Rössler A, Knabl L, von Laer D, Kimpel J. Neutralization Profile after Recovery from SARS-CoV-2 Omicron Infection. N Engl J Med 2022 (In eng). DOI: 10.1056/NEJMc2201607.

3. Carreno JM, Alshammary H, Tcheou J, et al. Activity of convalescent and vaccine serum against SARS-CoV-2 Omicron. Nature 2021. DOI: 10.1038/s41586-022-04399-5.

4. Cele S, Jackson L, Khoury DS, et al. Omicron extensively but incompletely escapes Pfizer BNT162b2 neutralization. Nature 2022;602(7898):654–656. (In eng). DOI: 10.1038/s41586-021-04387-1.

5. Tarke A, Coelho CH, Zhang Z, et al. SARS-CoV-2 vaccination induces immunological T cell memory able to cross-recognize variants from Alpha to Omicron. Cell 2022;185(5):847–859.e11. (In eng). DOI: 10.1016/j.cell.2022.01.015.

6. Keeton R, Tincho MB, Ngomti A, et al. T cell responses to SARS-CoV-2 spike cross-recognize Omicron. Nature 2022;603(7901):488–492. (In eng). DOI: 10.1038/s41586-022-04460-3.

7. GeurtsvanKessel CH, Geers D, Schmitz KS, et al. Divergent SARS-CoV-2 Omicron-reactive T and B cell responses in COVID-19 vaccine recipients. Sci Immunol 2022;7(69):eabo2202. (In eng). DOI: 10.1126/sciimmunol.abo2202.

8. Wratil PR, Stern M, Priller A, et al. Three exposures to the spike protein of SARS-CoV-2 by either infection or vaccination elicit superior neutralizing immunity to all variants of concern. Nat Med 2022;28(3):496–503. (In eng). DOI: 10.1038/s41591-022-01715-4.

9. Netzl A, Tureli S, LeGresley E, Mühlemann B, Wilks SH, Smith DJ. Analysis of SARS-CoV-2 Omicron Neutralization Data up to 2021-12-22. bioRxiv 2022:2021.12.31.474032. DOI: 10.1101/2021.12.31.474032.

10. Bruel T, Hadjadj J, Maes P, et al. Serum neutralization of SARS-CoV-2 Omicron sublineages BA.1 and BA.2 in patients receiving monoclonal antibodies. Nat Med 2022. DOI: 10.1038/s41591-022-01792-5.

11. Bowen JE, Sprouse KR, Walls AC, et al. Omicron BA.1 and BA.2 neutralizing activity elicited by a comprehensive panel of human vaccines. bioRxiv 2022:2022.03.15.484542. DOI: 10.1101/2022.03.15.484542.

12. Yu J, Collier A-rY, Rowe M, et al. Neutralization of the SARS-CoV-2 Omicron BA.1 and BA.2 Variants. New England Journal of Medicine 2022. DOI: 10.1056/NEJMc2201849.

13. Iketani S, Liu L, Guo Y, et al. Antibody evasion properties of SARS-CoV-2 Omicron sublineages. Nature 2022 (In eng). DOI: 10.1038/s41586-022-04594-4.

14. Wilks SH, Mühlemann B, Shen X, et al. Mapping SARS-CoV-2 antigenic relationships and serological responses. bioRxiv 2022:2022.01.28.477987. DOI: 10.1101/2022.01.28.477987.

15. van der Straten K, Guerra D, van Gils MJ, et al. Mapping the antigenic diversification of SARS-CoV-2. medRxiv 2022:2022.01.03.21268582. DOI: 10.1101/2022.01.03.21268582.

16. Neerukonda SN, Vassell R, Lusvarghi S, et al. SARS-CoV-2 Delta Variant Displays Moderate Resistance to Neutralizing Antibodies and Spike Protein Properties of Higher Soluble ACE2 Sensitivity, Enhanced Cleavage and Fusogenic Activity. Viruses 2021;13(12). DOI: 10.3390/v13122485.

17. Hu Y-F, Hu J-C, Gong H-R, et al. Computation of Antigenicity Predicts SARS-CoV-2 Vaccine Breakthrough Variants. Frontiers in Immunology 2022;13 (Original Research) (In English). DOI: 10.3389/fimmu.2022.861050.

18. Mykytyn AZ, Rissmann M, Kok A, et al. Omicron BA.1 and BA.2 are antigenically distinct SARS-CoV-2 variants. bioRxiv 2022:2022.02.23.481644. DOI: 10.1101/2022.02.23.481644.

19. Riepler L, Rössler A, Falch A, et al. Comparison of Four SARS-CoV-2 Neutralization Assays. Vaccines (Basel) 2020;9(1) (In eng). DOI: 10.3390/vaccines9010013.

20. Smith DJ, Lapedes AS, de Jong JC, et al. Mapping the antigenic and genetic evolution of influenza virus. Science 2004;305(5682):371–6. (In eng). DOI: 10.1126/science.1097211.

21. Wilks S. Racmacs: R Antigenic Cartography Macros. (https://acorg.github.io/Racmacs/index.html).

22. Fonville JM, Wilks SH, James SL, et al. Antibody landscapes after influenza virus infection or vaccination. Science 2014;346(6212):996–1000. (In eng). DOI: 10.1126/science.1256427.

23. R Core Team. R: A Language and Environment for Statistical Computing. R Foundation for Statistical Computing. (https://www.R-project.org/).

24. Knabl L, Mitra T, Kimpel J, et al. High SARS-CoV-2 seroprevalence in children and adults in the Austrian ski resort of Ischgl. Commun Med (Lond) 2021;1(1):4. (In eng). DOI: 10.1038/s43856-021-00007-1.

25. Bekliz M, Adea K, Vetter P, et al. Neutralization of ancestral SARS-CoV-2 and variants Alpha, Beta, Gamma, Delta, Zeta and Omicron by mRNA vaccination and infection-derived immunity through homologous and heterologous variants. medRxiv 2022:2021.12.28.21268491. DOI: 10.1101/2021.12.28.21268491.

26. Khan K, Karim F, Cele S, et al. Omicron infection of vaccinated individuals enhances neutralizing immunity against the Delta variant. medRxiv 2022:2021.12.27.21268439. DOI: 10.1101/2021.12.27.21268439.

27. Seaman MS, Siedner MJ, Boucau J, et al. Vaccine Breakthrough Infection with the SARS-CoV-2 Delta or Omicron (BA.1) Variant Leads to Distinct Profiles of Neutralizing Antibody Responses. medRxiv 2022:2022.03.02.22271731. DOI: 10.1101/2022.03.02.22271731.

28. Corbett KS, Gagne M, Wagner DA, et al. Protection against SARS-CoV-2 Beta variant in mRNA-1273 vaccine–boosted nonhuman primates. Science 2021;374(6573):1343–1353. DOI: doi:10.1126/science.abl8912.

29. Cameroni E, Bowen JE, Rosen LE, et al. Broadly neutralizing antibodies overcome SARS-CoV-2 Omicron antigenic shift. Nature 2022;602(7898):664–670. DOI: 10.1038/s41586-021-04386-2.

30. Zhou H, Tada T, Dcosta BM, Landau NR. Neutralization of SARS-CoV-2 Omicron BA.2 by Therapeutic Monoclonal Antibodies. bioRxiv 2022:2022.02.15.480166. DOI: 10.1101/2022.02.15.480166.

31. Bruel T, Hadjadj J, Maes P, et al. Serum neutralization of SARS-CoV-2 Omicron sublineages BA.1 and BA.2 in patients receiving monoclonal antibodies. Nature Medicine 2022. DOI: 10.1038/s41591-022-01792-5.

