## Supplementary Appendix for "BA.2 omicron differs immunologically from both BA.1 omicron and pre-omicron variants"

**List of Investigators**

Annika Rössler[1,*]

Antonia Netzl[2,*]

Ludwig Knabl[3]

Helena Schäfer[1]

Samuel H. Wilks[2]

David Bante[1]

Barbara Falkensammer[1]

Wegene Borena[1]

Dorothee von Laer[1]

Derek Smith[2,#]

Janine Kimpel[1,#]

[1]Institute of Virology, Department of Hygiene, Microbiology and Public Health, Medical University of Innsbruck, Peter-Mayr-Str. 4b, 6020 Innsbruck, Austria

[2]University of Cambridge, Center for Pathogen Evolution, Department of Zoology, Cambridge, UK

[3]Tyrolpath Obrist Brunhuber GmbH, Hauptplatz 4, 6511, Zams, Austria

*Contributed equally

[#]Corresponding authors:

Derek Smith and Janine Kimpel

**Supplementary Methods**

*Patient Characteristics*

Patients were divided into three groups depending on their numbers of exposure to infection/vaccination. Single exposure was defined as unvaccinated with a single infection (n=10 infected during first wave (wild type), n=10 alpha, n=8 beta, n=7 delta, n=18 BA.1 omicron, and n=12 BA.2 omicron. Double exposure was defined as either unvaccinated individuals with two subsequent infections (n=15 with pre-omicron infection followed by BA.1 re-infection and n=3 with pre-omicron infection followed by BA.2 re-infection) or two vaccine doses without any history of infection (two homologous doses of mRNA-1273 (n=10), ChAdOx-S1 (n=10) or BNT162b2 (n=11) respectively or heterologous vaccination with ChAdOX-S1 as first and BNT162b2 as second dose (n=10)). Three or more exposures were defined as vaccinated individuals with breakthrough infections (n=6 with delta breakthrough after two doses of ChAdOx-S1, n=22 with delta breakthrough after two doses of BNT162b2, n=14 with two or three vaccine doses and BA.1 omicron breakthrough, and n=8 with two or three vaccine doses and BA.2 omicron breakthrough) or vaccinated individuals with a previous pre-omicron breakthrough followed by a BA.1 omicron breakthrough (n=11).

*Map Diagnostics*

Map diagnostic tools were used to assess the quality of the antigenic map with P.1.1 reactivity adjusted by -1 on the $\log_2$ scale (Figure 2, Supplementary Figure S2).

*Map Dimensionality*

First, a dimensionality test was performed with *Racmacs'* [1] *dimensionTestMap* function in dimensions 1 to 5 with a test proportion of 0.1, the minimum column basis set to "none", the fixed column bases of the current map, and 100 replicates per dimension with 1000 optimizations per replicate. The Root Mean Square Error RMSE between measured and predicted titers was used to judge which dimensionality best represents the antigenic relationships of the titer data. Figure S3 shows that the error between predicted and measured titers is smallest in two dimensions, indicating that the titer data is best represented by a 2D map.

*Correlation of fitted and measured titers*

Next, the correlation of fitted and measured titers of the map was assessed by converting map distances into $\log_2$ titers. As one antigenic unit in the map corresponds to one two-fold dilution in the neutralization assay, subtracting the Euclidean distance for each serum-antigen pair from the maximum $\log_2$ titer of the specific serum gives $\log_2$ titers.

Doing this for each serum group revealed good correlation of measured and fitted $\log_2$ titers (Figure S4). In the omicron convalescent serum groups, measured titers exceeded fitted titers slightly. A large number of below detectable titers, however, impairs the accurate positioning of a serum and explains the larger difference between measured and fitted titers in the omicron convalescent compared to other serum groups.

*Map robustness*

Bootstrapping on sera and antigens was performed to determine how robust the map is to measurement noise and the exclusion of samples and antigen variants. *Racmacs'* [1] *bootstrapMap* function with the "noisy" method was used to simulate measurement noise, doing 1000 repeats and 100 optimizations per repeat, bootstrapping both sera and antigens with normally distributed measurement noise with a standard deviation of 0.7 added to either the titer, the antigen or both (Figure S5). The same function and optimization routine was used to test the impact of serum and antigen exclusion on the map, using the "resample" method with antigen and titer noise standard deviation of 0.7, and resampling only antigens, sera or resampling both (Figure S6). To test the exclusion of entire serum groups (Figure S7) or antigen variants (Figure S8), maps were constructed excluding the specific sera or variant with 500 optimizations per map and the minimum column basis set to "none".

Adding noise to antigen reactivity adds noise to all titers measured against the specific antigen, whereas titer noise adds noise to a single measurement. The effects of which are shown in Figure S5. The variants' positions were robust to titer noise (Figure S5B). The positions of non-omicron variants varied by one to two antigenic units when adding noise to antigenic reactivity (Figure S5 A, C). Antigen noise resulted in hemishering of BA.1 omicron, referring to two distinct optima for the variant's position. The big effect on variant position by some noise can be understood if the added noise moves an originally detectable titer below the detection threshold, or vice-versa. In case of the two omicron sub-lineages, many titers were at the border or below detectable, hence adding artificial measurement noise affected the positioning of these variants more than the other variants.

This can also be seen in Figure S6, where 1000 optimization runs were performed in which both antigens and sera were randomly resampled with replacement (Figure S6A), only antigen variants were resampled (Figure S6B) or individual serum samples were randomly sampled with replacement (Figure S6C). Resampling variants had little effect on non-omicron variant positioning, but again two different optima were found for BA.1 and BA.2 omicron. While serum bootstrapping had little effect on both variants and sera (Figure S6C), variant bootstrapping demonstrated the uncertainty of BA.1 and BA.2 omicron convalescent sera positioning (Fig. S6 B), which had detectable titers against their root variant but very low to non-detectable titers against other variants.

However, the distance of BA.1 and BA.2 omicron from vaccine sera and to each other remained stable.

To investigate which serum groups or variants had a particularly large impact on map topology, maps excluding single serum groups (Figure S7) or antigen variants (Figure S8) were constructed. Excluding BA.1 omicron conv. sera resulted in the effect on BA.1 and BA.2 omicron positions we observed when adding noise or resampling variants in map construction. Removal of other serum groups had no effect on variant positions.

Excluding the D614G and beta variant had a similar effect and moved both omicron variants to a second optimal position (Figure S8).

*Map cross-validation*

For cross validation, *Racmacs'* [1] *dimensionTestMap* function was employed to optimize maps in 2 dimensions with a test proportion of 0.1. 1000 replicates and 500 optimizations per replicate were performed and the difference between measured and predicted titer calculated on the $\log_2$ scale.

Briefly, 1000 optimizations runs were performed in which only 90% of titers were used for map construction and the remaining 10% predicted by subtracting the Euclidean map distance for each serum-antigen pair from the maximum $log_2$ titer of the specific serum. The mean difference between measured and predicted $log_2$ titer was less one two-fold for detectable titers (Figure S9). However, this differed largely across serum groups and antigen variants (Figure S10). The variability of BA.1 and BA.2 omicron conv. sera positions, and BA.1 and BA.2 omicron variant positions seen in the bootstrap map resulted in much lower predicted than measured titers for these variants in the specific serum groups. As before, if detectable titers against these variants were excluded from the map creation, their position became inaccurate and hence sera with only detectable titers against BA.1 or BA.2 omicron, which was true for the majority of BA.1 and BA.2 omicron conv. sera, could not be positioned either.

*Individual antibody landscapes*

Antibody landscapes [2] were constructed as described in *Materials & Methods.* With the map shown in Figure 2 serving as base plane, antibody levels for each serum were plotted in a third dimension and a continuous surface fitted to the available titer data. The reactivity profile of serum groups exposed to a single variant, such as the convalescent and double vaccinated serum groups (Figure S11 A-J), was assumed to adopt a cone like shape, with the highest point at the specific serum coordinate and a constant decrease across antigenic space of one two-fold per antigenic unit.[3] This assumption does not apply to serum groups exposed to more than one variant, as high titers against both encountered variants are expected. For these groups (Figure S11 K-Q), each serum's coordinates were fitted to optimize the serum's position in the map shown in Figure 2 and the surface's slope was fitted per serum group, as the decrease of reactivity was assumed to be the same for individuals with the same exposure history. The GMT landscapes show the average of all individual antibody landscapes.

The vaccine landscapes (Figure S11 A-D) were the most homogeneous in terms of titer magnitudes within the serum group. Further, the different vaccine regiments elicted landscapes similar to each other. The individual antibody profiles of most most convalescent groups were homogenous. With the exception of one delta serum with high P.1.1 titers and two BA.2 conv. sera, all had highest titers against their non-homologous variant. Antibody profiles such as these could be due to unknown prior infections. The specific delta sample, however, was a PCR-confirmed first infection, suggesting a different reason for the high titer against P.1.1. To investigate potential prior infection of the two cross-reactive BA.2 omicron conv. samples, maps were created without these two sera and antibody landscapes fitted to their titers (Figure S12). The two sera did not impact the variant positions in the map (Figure S12 A-C). Their antibody profiles, however, looked more similar to BA.2 omicron breakthrough infections (sample G780) or reinfections (sample G776) than to BA.2 omicron first infections (Figure S12 D). As the map without those two sera did not differ from the map including the sera, the general conclusion of BA.2 omicron being positioned between pre-omicron variants and BA.1 omicron did not change.

The antibody landscapes after delta breakthrough were similar in both magnitude and shape independent of Astra Zeneca or Pfizer vaccination (Figure S11 K-L), and were further comparable to BA.1 and BA.2 omicron breakthrough infections. Breakthrough landscapes after either omicron sub-lineage infection or vaccinated with prior infection and BA.1 omicron breakthrough were almost indistinguishable both in titer magnitudes and landscape slope (Figure S11 M-P). All multi-exposure landscapes, including BA.2 omicron reinfected landscapes (Figure S11 Q), exhibited a broad reactivity profile, suggesting high degrees of cross-reactivity and good protection against variants within the current mapped antigenic space.

The largest variation of individual landscapes was found in the BA.1 omicron breakthrough and reinfected serum groups (Figure S11 M, O). To investigate the impact of number of vaccinations and variant of prior infection, these two groups were split into subgroups and antibody landscapes were constructed for each subgroup (Figure S13). This revealed that the landscape shape of double and triple vaccinated individuals with BA.1 omicron breakthrough infection was almost identical, and the observed variation was due to generally higher titers after three vaccine doses (Figure S13 A-B). The shapes of reinfection landscapes, however, differed depending on the first encountered variant (Figure S13 C-D). Prior infection with D614G resulted in antibody profiles similar to the vaccinated and infected, and previous infection with vaccination groups, with reactivity peaking in the area of D614G and slowly decreasing towards the omicron variants. Sera infected with delta followed by BA.1 omicron had lower titers in the upper area of the map (D614G, alpha, beta), with individual landscapes having higher BA.1 omicron titers than delta titers, resulting in a landscape shape almost mirroring the D614G landscape. Only one sample was available with a prior alpha infection, which exhibited very low titers against BA.1 omicron and much less cross-reactivity than the other BA.1 reinfection landscape.

**Supplementary Figures**

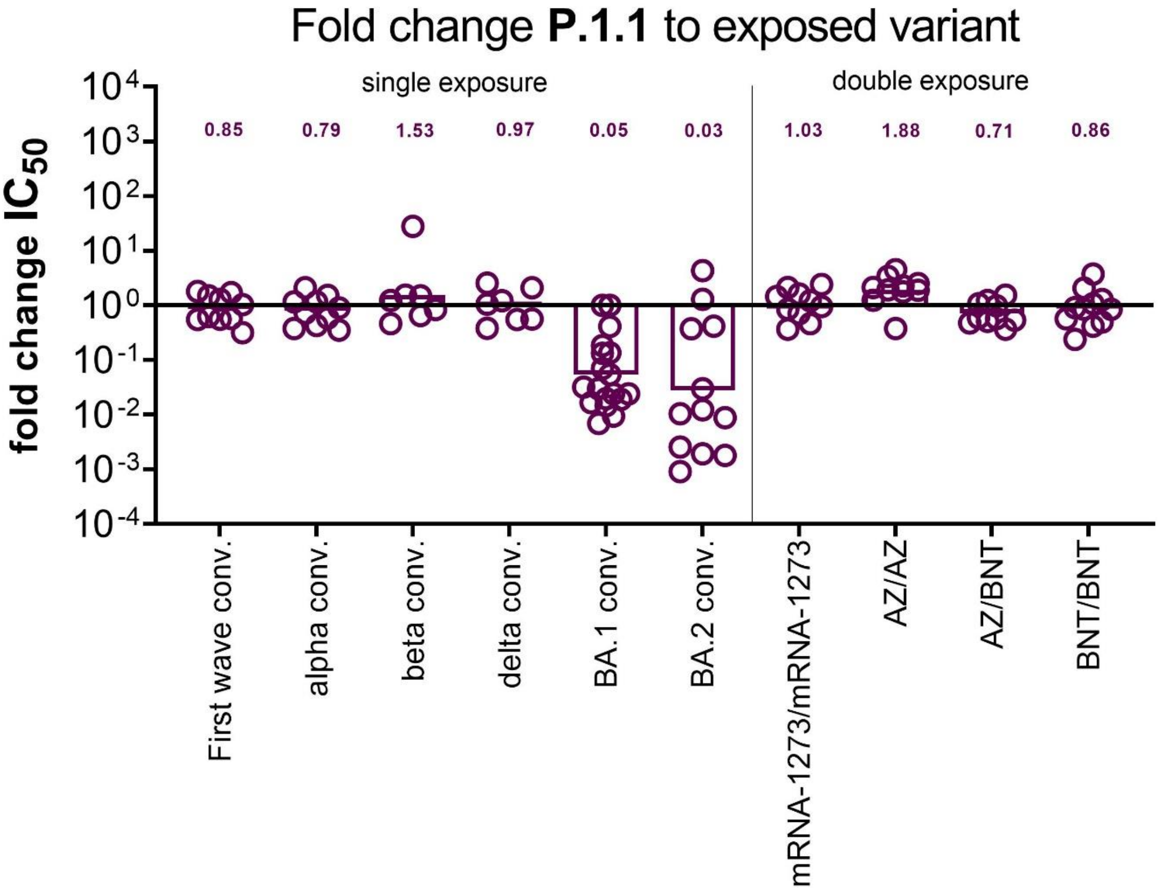

**Figure S1. Fold change neutralizing antibody titers gamma versus exposed variant.** For each sample, the fold change titer in neutralizing antibodies ($IC_{50}$) between gamma and the exposed variant was calculated. For first wave convalescent or two dose vaccinated individuals the D614G titers were used as exposed variant. Shown are individual samples and geometric mean change as bars. Numbers indicate the geometric mean change for each group.

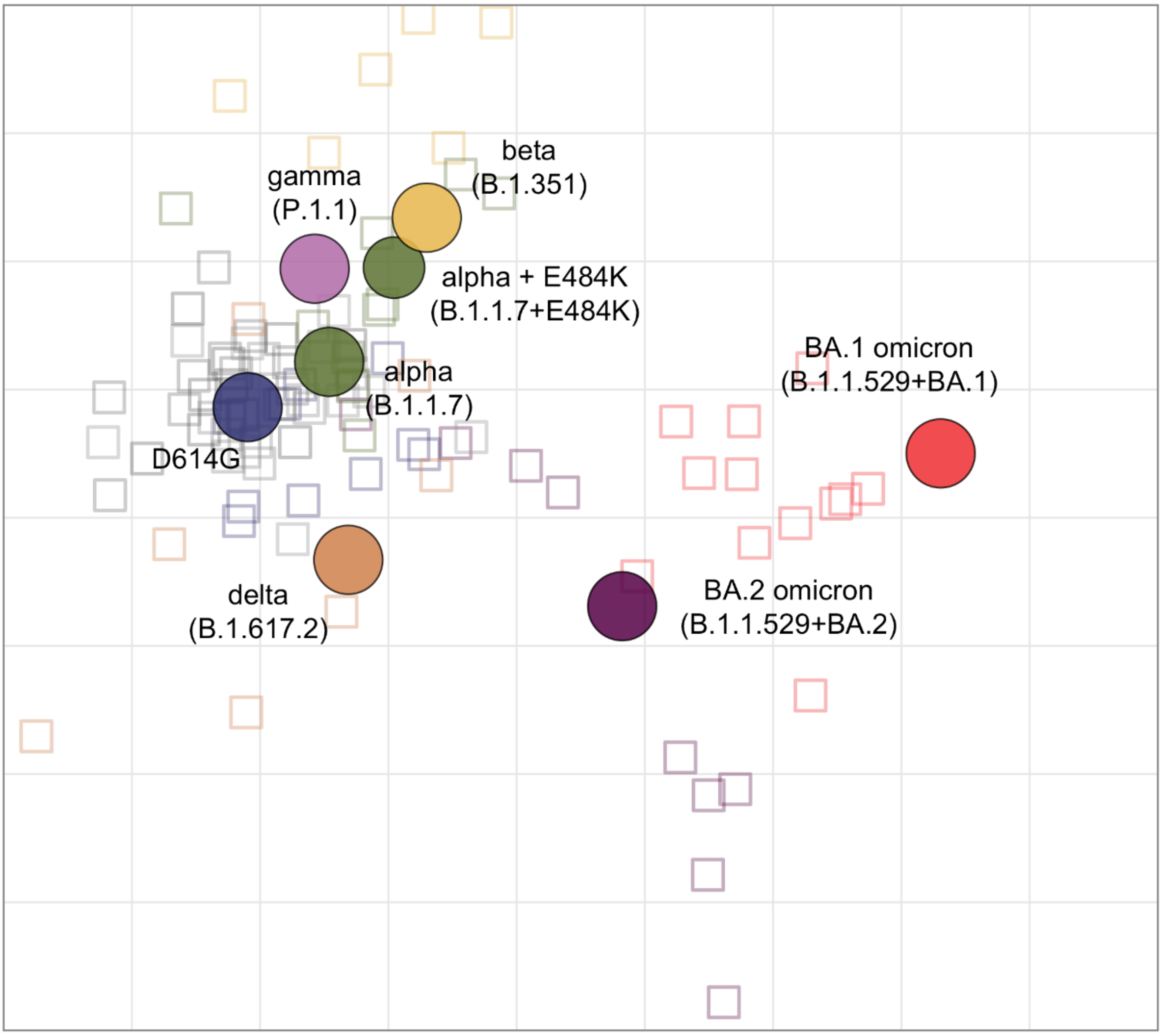

**Figure S2. Non-zoomed in version of the antigenic map.** Virus variants are shown as colored circles, sera as open squares with the color corresponding to the infecting variant. Vaccine sera are shown in grey tones (from dark to light: mRNA-1273/mRNA-1273, BNT162b2/BNT162b2, ChAdOx-S1/BNT162b2, ChAdOx-S1/ChAdOx-S1). The alpha + E484K variant is shown as smaller circle due to its additional substitution compared to the alpha variant. The x- and y-axis represent antigenic distances with one grid square corresponding to one two-fold serum dilution of the neutralization titer. The map orientation within x- and y-axis is free as only relative distances can be inferred.

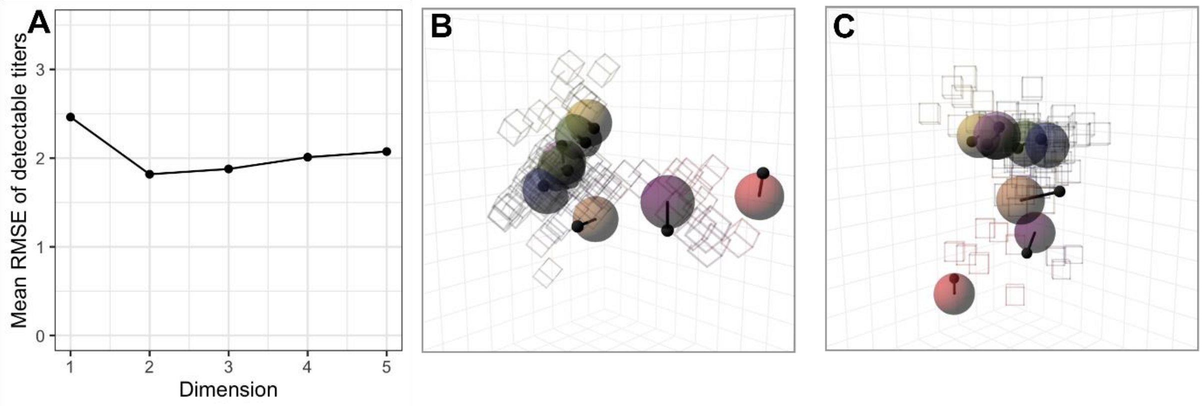

**Figure S3. Map dimensionality. A** Dimensionality test: RMSE between map and measured titers for detectable titers in 1 to 5 dimensions. Per dimension, 100 map replicates were constructed from 90% of measured titers with 1000 optimizations per replicate. The titers of the remaining 10% were predicted in each run and the RMSE calculated by comparing the predicted to the measured titers on the $log_2$ scale. **B, C** Side and front view of the map optimized in 3 dimensions with arrows pointing to the variants' position in the 2D map.

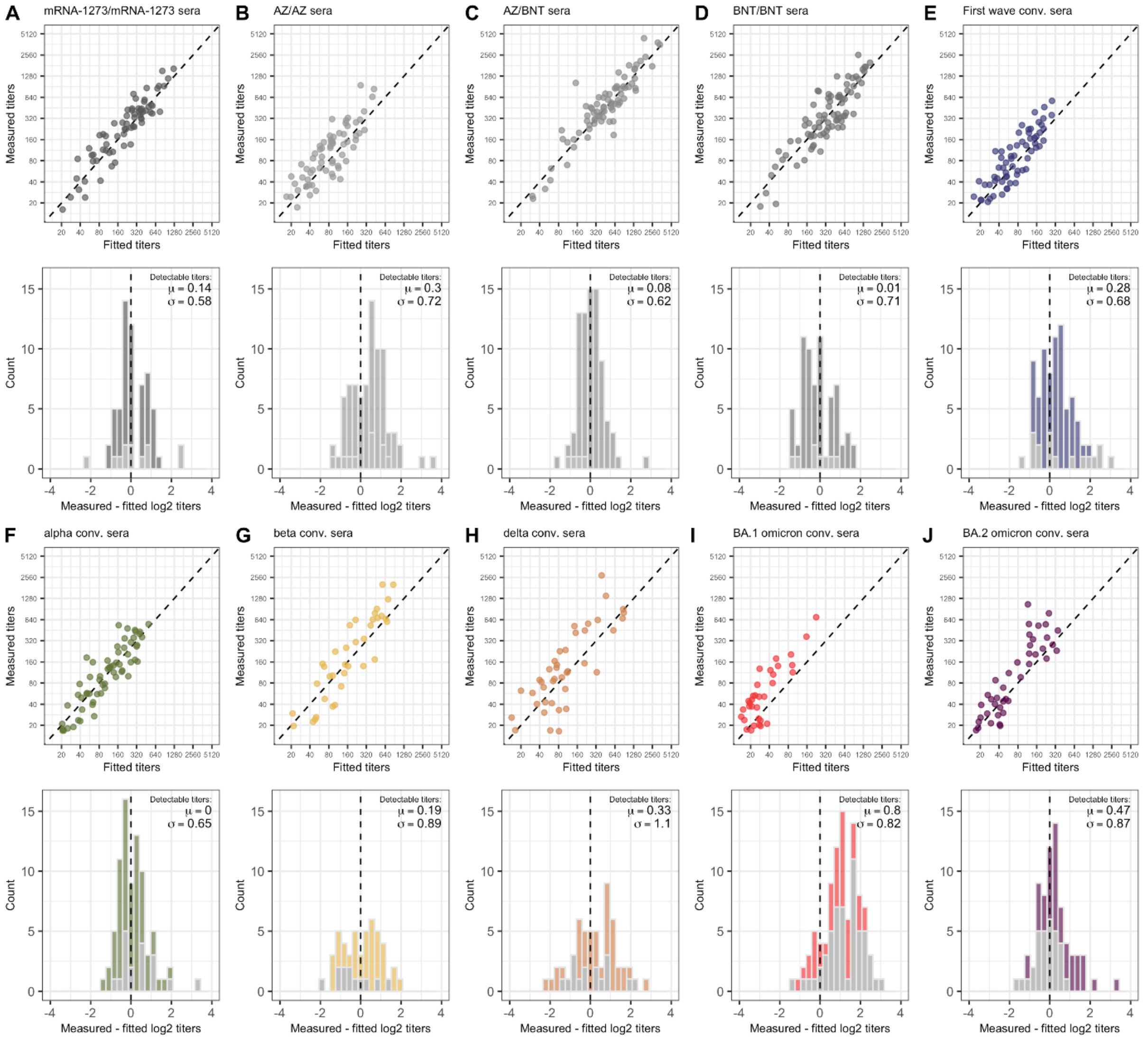

**Figure S4. Goodness of map fit per serum group.** The top panels show the correlation of detectable measured and fitted titers in the 2D map with P.1.1 reactivity adjustment. Measured P.1.1 titers were reduced by one two-fold to match the reactivity adjustment in the map. Map distances were converted into $\log_2$ titers by subtracting the Euclidean distance for each serum-antigen pair from the maximum $\log_2$ titer of the specific serum. The bottom panels show the residuals of measured against fitted titers on the $\log_2$ scale, light grey marks pairs with the measured titer below the assay detection threshold. The mean and mean-centered standard deviation of differences between fitted and detectable measured titers are given in the legend of each bottom row panel. This was done for the serum groups used to construct the map: **A** mRNA-1273/mRNA-1273, **B** ChAdOx-S1/ChAdOx-S1, **C** ChAdOx-S1/BNT162b2, **D** BNT162b2/BNT162b2, **E** first wave conv., **F** alpha conv., **G** beta conv., **H** delta conv., **I** BA.1 omicron conv., **J** BA.2 omicron conv.

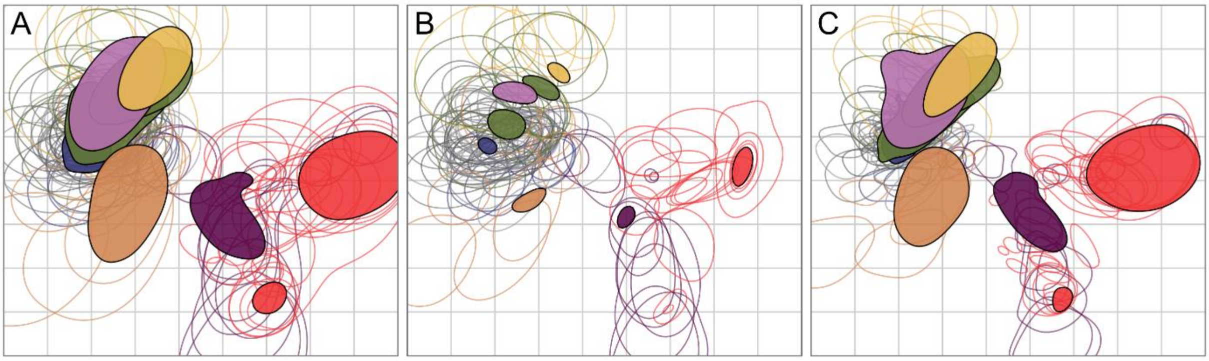

**Figure S5. Assessing map robustness to measurement uncertainty by bootstrapping.** 1000 bootstrap repeats were performed with 100 optimizations per repeat. Normally distributed measurement noise with a standard deviation of 0.7 was added to **A** titers and antigen reactivity, **B** only titers, and **C** only antigen reactivity. The colored regions mark 68% (one standard deviation) of the positional variation for each variant (filled shapes) and sera (open shapes). The colors correspond to the colors used in Figure 2.

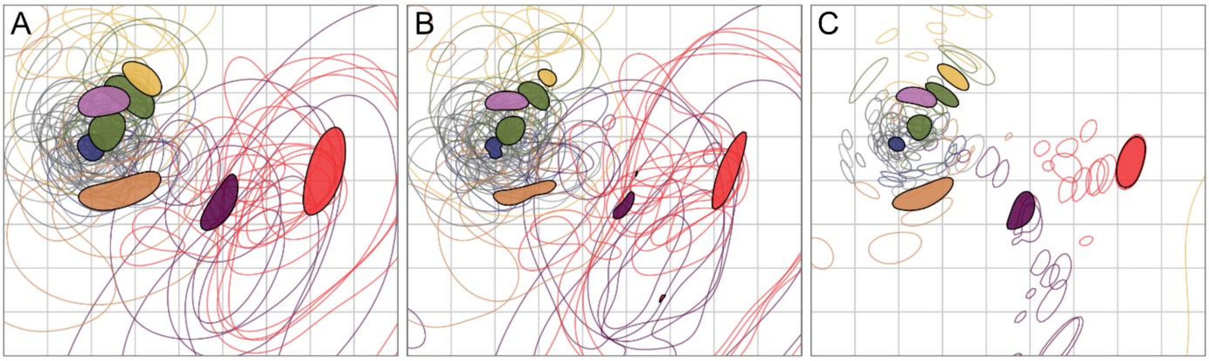

**Figure S6. Assessing map robustness to the exclusion of measurements by bootstrapping.** 1000 bootstrap repeats were performed with 100 optimizations per repeat. For each repeat, a random subset of titer measurements was taken with replacement. The bootstrapping was performed on **A** variants and sera, **B** only variants, and **C** only sera. The colored regions mark 68% (one standard deviation) of the positional variation for each variant (filled shapes) and sera (open shapes). The colors correspond to the colors used in Figure 2.

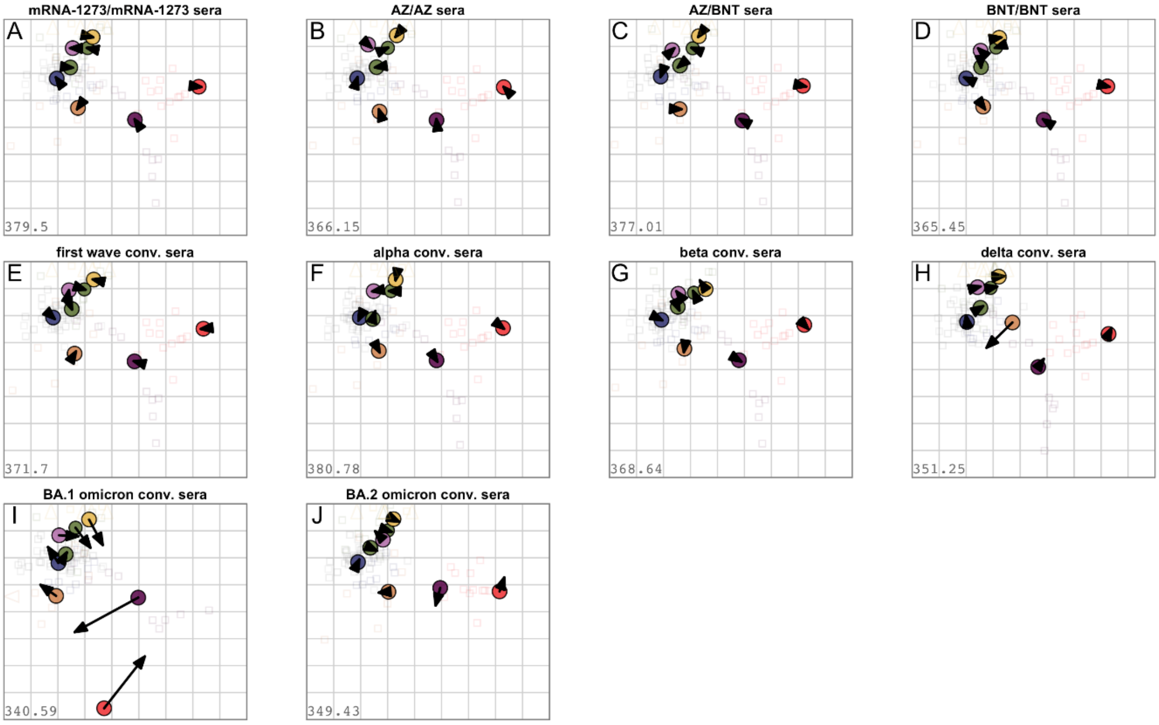

**Figure S7. Assessing map robustness to the exclusion of sera.** Each serum group was removed and the map reoptimized. Arrows point to the position of each variant in the map shown in Figure 2, for color correspondence refer to this map. A small arrow length indicates similar variant positions and map robustness to the exclusion of the particular serum group. Triangles point to sera positioned outside the plotting area. Maps without **A** mRNA-1273/mRNA-1273, **B** ChAdOx-S1/ChAdOx-S1, **C** ChAdOx-S1/BNT162b2, **D** BNT162b2/BNT162b2, **E** first wave conv., **F** alpha conv., **G** beta conv., **H** delta conv., **I** BA.1 omicron conv., **J** BA.2 omicron conv.

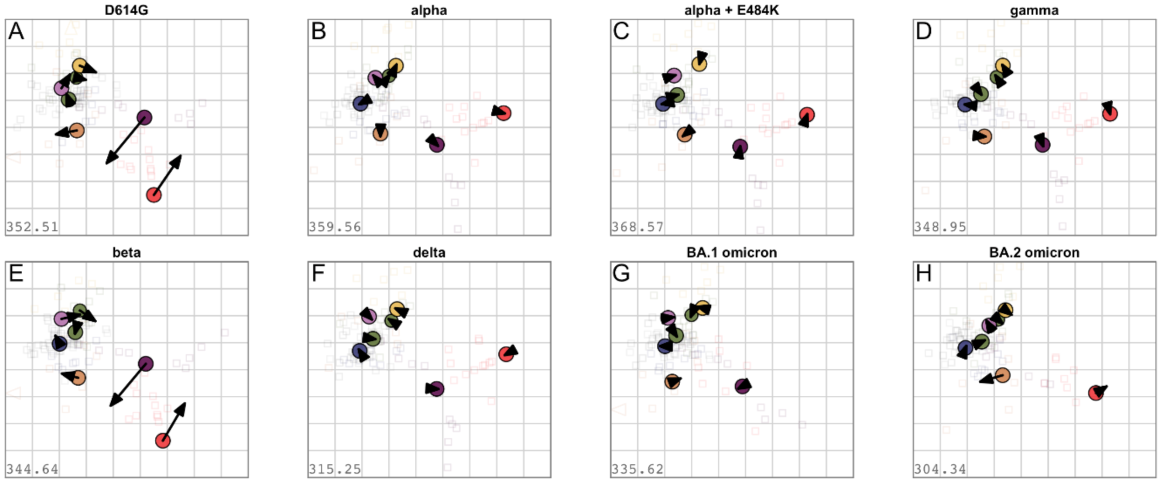

**Figure S8. Assessing map robustness to the exclusion of antigen variant.** Each antigen variant was removed and the map reoptimized. Arrows point to the position of each variant in the map shown in Figure 2, for color correspondence refer to this map. A small arrow length indicates similar variant positions and robustness to the exclusion of the particular antigen variant. Triangles point to sera positioned outside the plotting area. Maps without **A** D614G, **B** alpha (B.1.1.7), **C** alpha + E484K (B.1.1.7 + E484K), **D** gamma (P.1.1), **E** beta (B.1.351), **F** delta (B.1.617.2), **G** BA.1 omicron (B.1.1.529+BA.1), **H** BA.2 omicron (B.1.1.529+BA.2)**.**

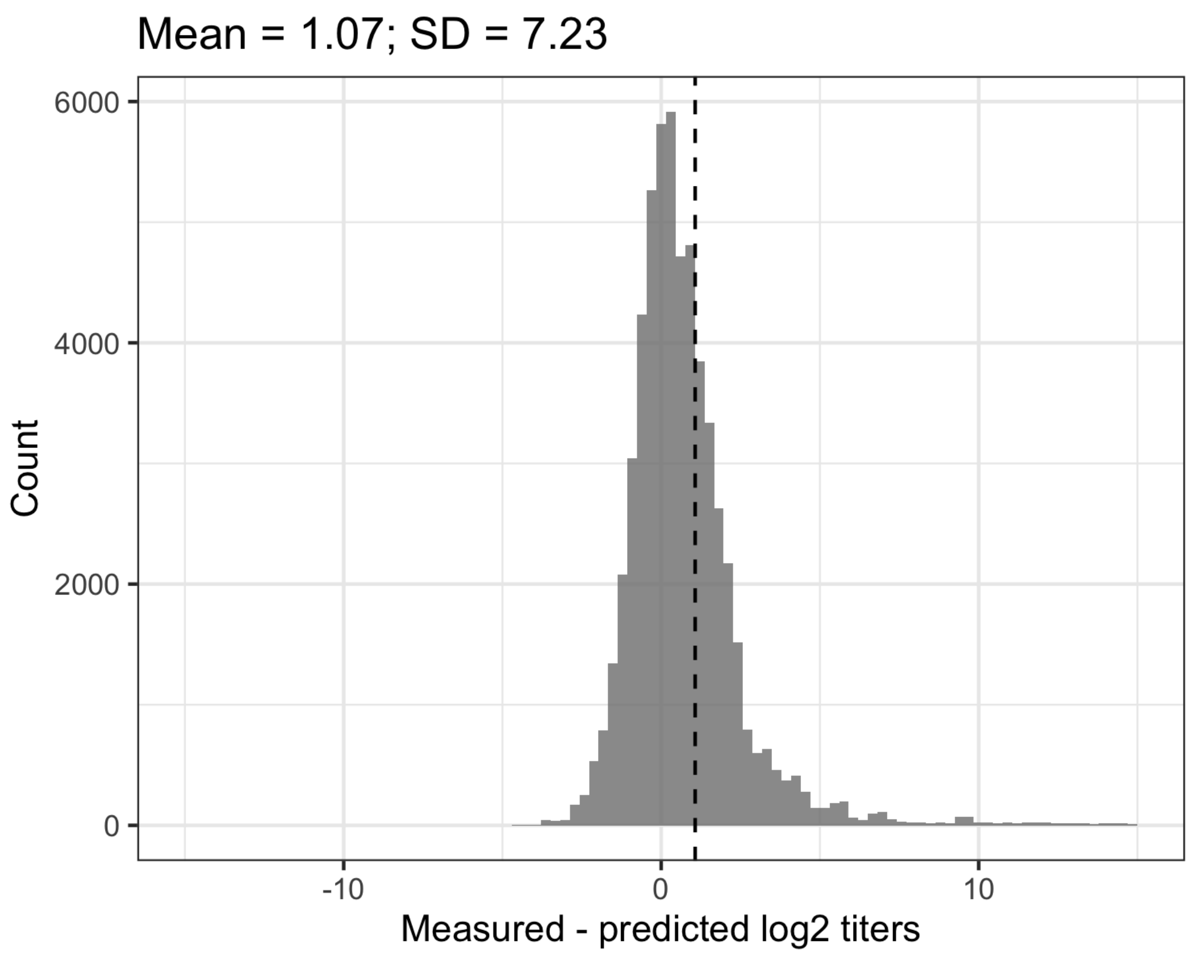

**Figure S9. Map cross-validation residual titers.** 1000 optimization runs were performed with only 90% of measured titers used for map construction by artificially masking 10% of measurements. The missing $log_2$ titers were predicted by subtracting the Euclidean map distance for each serum-antigen pair from the maximum $log_2$ titer of the specific serum. The difference between predicted and detectable measured titers on the $log_2$ scale was calculated, the mean is indicated by the dashed line. The mean and mean-centered standard deviation are given. The upper x-axis limit has been set to 15 for plotting purposes, very few residuals in the BA.1 and BA.2 omicron convalescent groups were larger than 100, due to inaccurate positioning of sera.

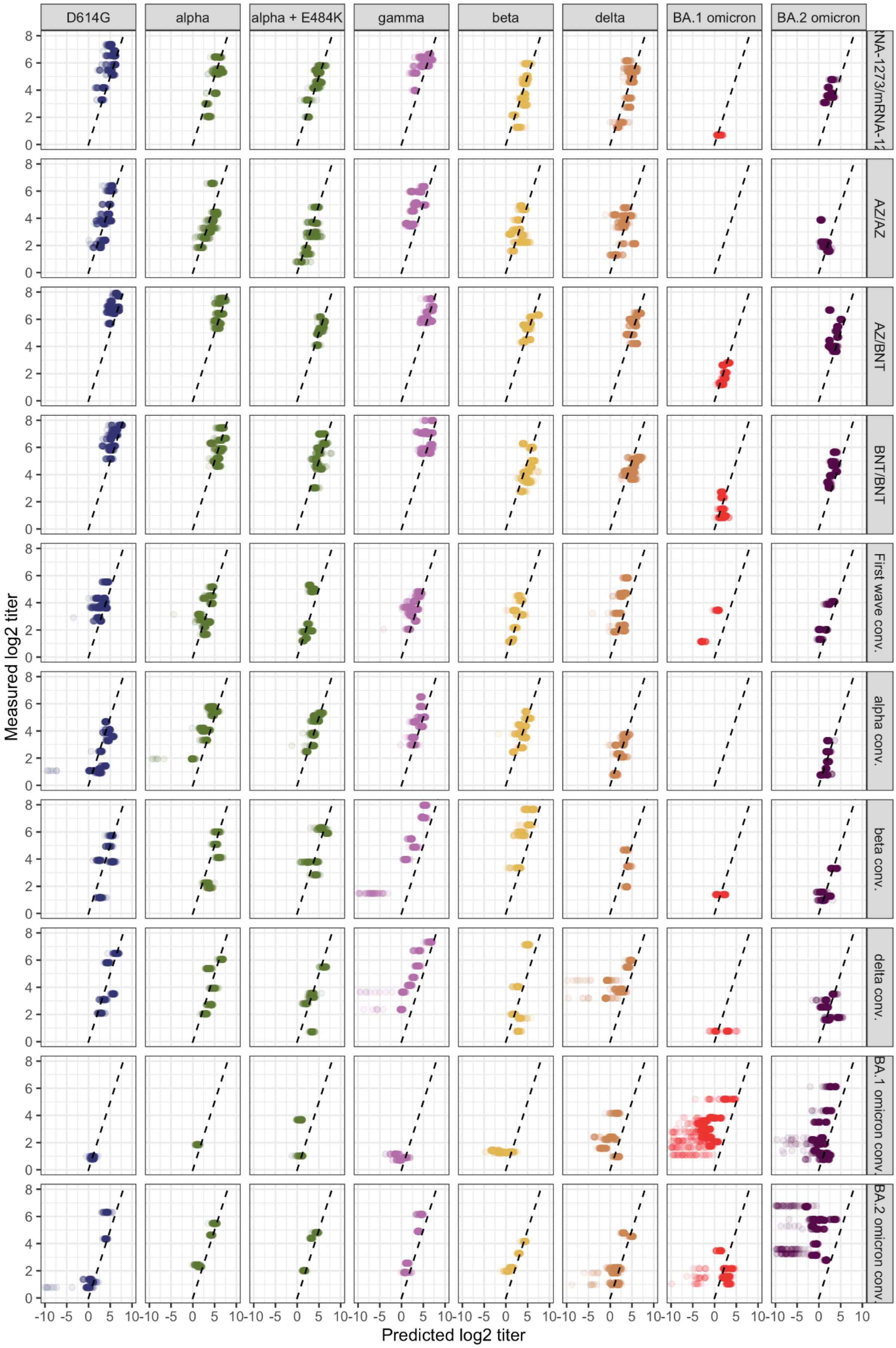

***Figure S10: Map cross-validation predicted vs. measured titers.*** *Figure caption on next page.*

**Figure S10. Map cross-validation predicted vs. measured titers.** 1000 optimization runs were performed with only 90% of measured titers used for map construction by artificially masking 10% of measurements. The missing $\log_2$ titers were predicted by subtracting the Euclidean map distance for each serum-antigen pair from the maximum $\log_2$ titer of the specific serum. The detectable measured over predicted log2 titers are shown per serum group and antigen variant. No BA.1 omicron titers were above the detection threshold in the alpha convalescent and AZ/AZ vaccinated serum groups. The lower x-axis limit has been set to -10 for plotting purposes, very few residuals in the BA.1 and BA.2 omicron convalescent groups were larger than 100, due to inaccurate positioning of sera. Many thresholded titers in the original dataset for the omicron variants and in the omicron serum group resulted in inaccurate predictions if the detectable measurements were masked in map making.

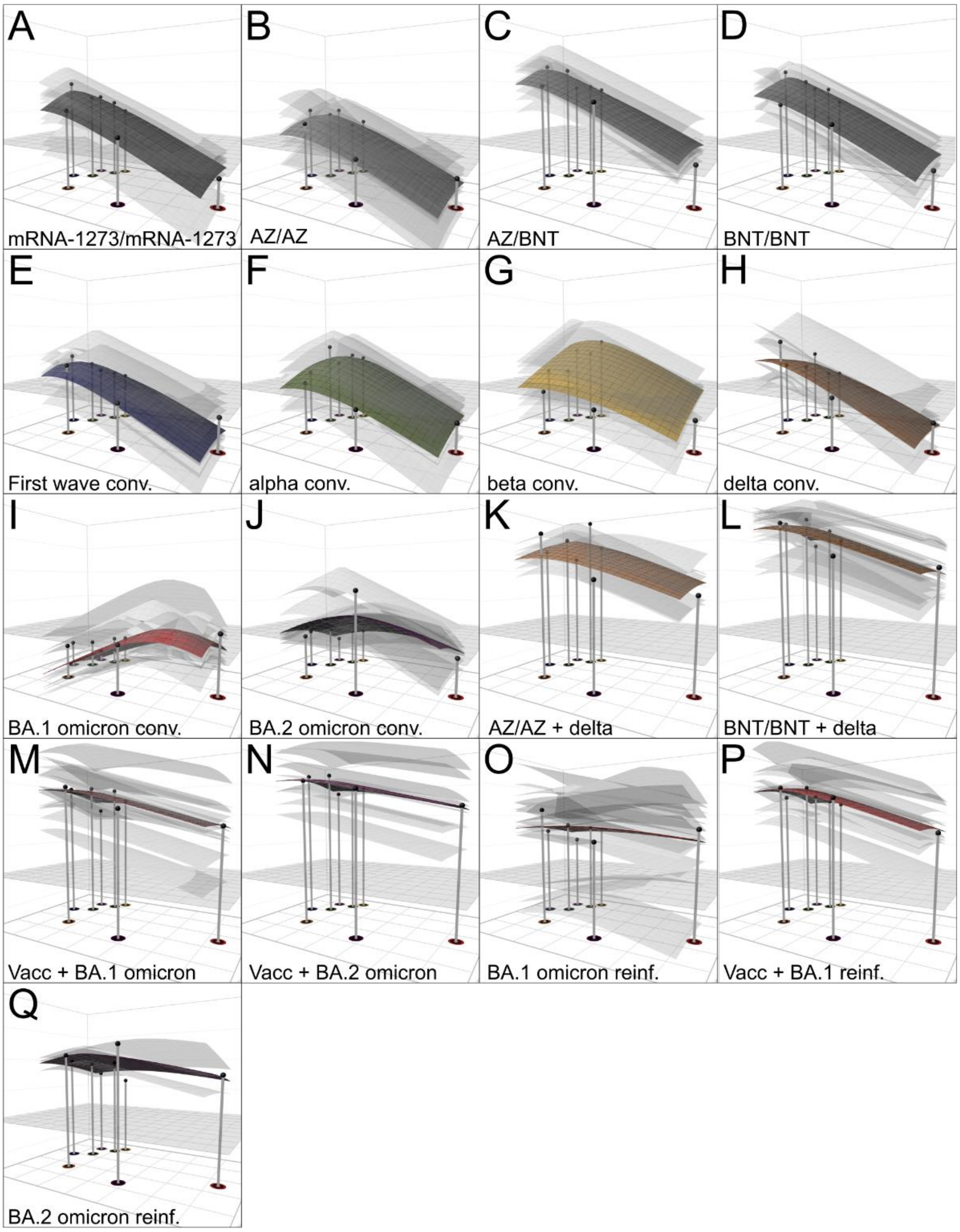

**Figure S11. Individual serum antibody landscapes.** The colored surfaces represent the GMT antibody landscapes of the different serum groups, the grey transparent surfaces individual serum landscapes. Serum reactivity bias was adjusted for GMT calculation as described in (Wilks et al., 2022). The map shown in Figure 2 serves as base plane, the z-axis reflects $\log_2$ titers with each two-fold increase marked starting from titer 20. The grey plane at titer 50 serves as reference. The impulses show serum group GMTs for each variant. The landscapes are shown for the following serum groups: **A** mRNA-1273/mRNA-1273, **B** ChAdOx-S1/ChAdOx-S1, **C** ChAdOx-S1/BNT162b2, **D** BNT162b2/BNT162b2, **E** first wave conv., **F** alpha conv., **G** beta conv., **H** delta conv., **I** BA.1 omicron conv., **J** BA.2 omicron conv., **K** AZ/AZ + delta breakthrough, **L** BNT/BNT + delta breakthrough, **M** BA.1 omicron breakthrough, **N** BA.2 omicron breakthrough, **O** BA.1 omicron reinfection, **P** Vaccinated, previous non-omicron infection and BA.1 omicron infection, **Q** BA.2 omicron reinfection. Serum coordinates and landscapes for **K-Q** were fitted as described in *Materials & Methods.*

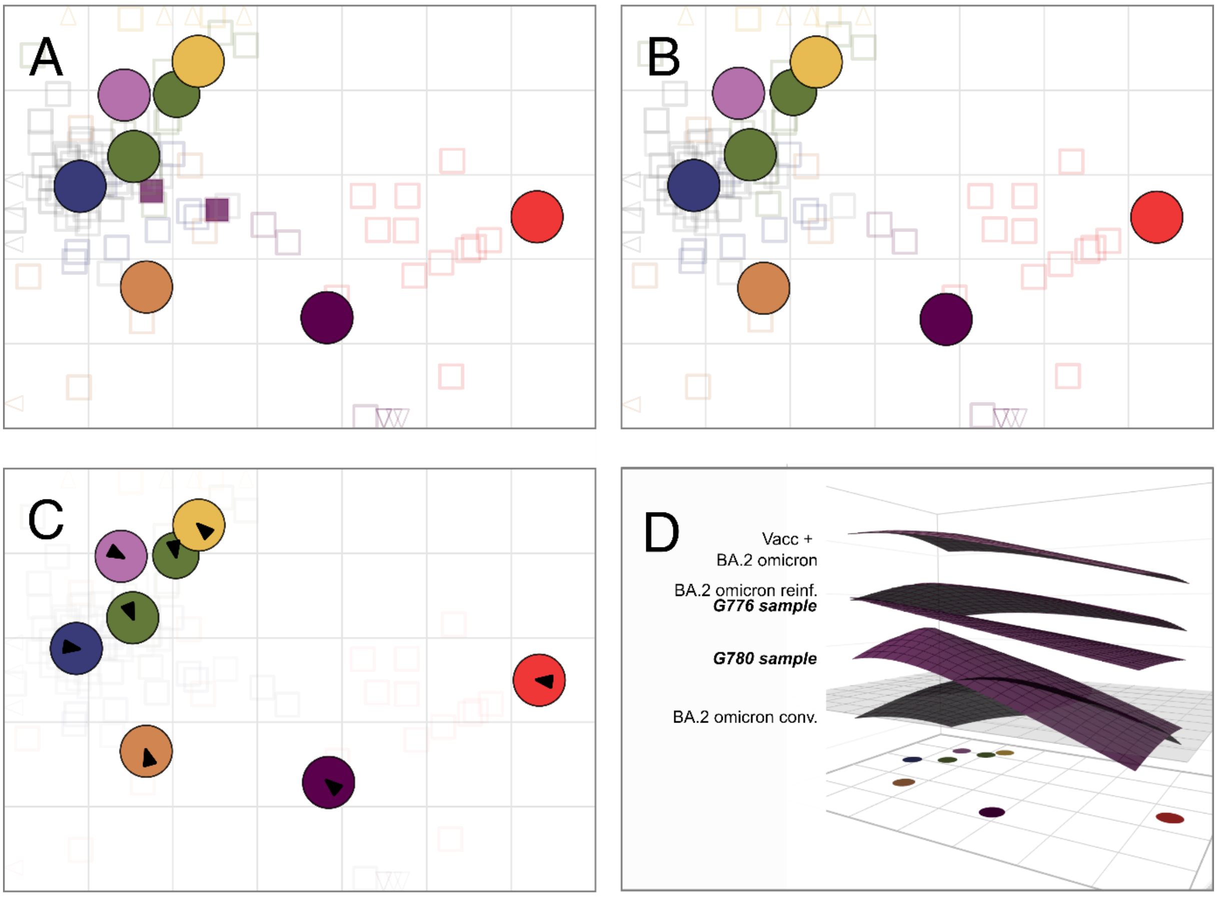

**Figure S12. Testing effect of cross-reactive BA.2 omicron convalescent sera on map and landscape.** **A** Map in Figure 2 shown with two cross-reactive BA.2 convalescent sera (G778, G780) highlighted in dark purple. **B** A map was constructed excluding these two sera to test their effect on variant positioning. **C** Map in **B** with arrows pointing to the variant's position in map **A.** The small arrows indicate that the two cross-reactive samples do not impact BA.2 positioning. **D** GMT Antibody landscapes of serum groups which encountered BA.2 omicron (Vacc + BA.2 omicron breakthroughs, BA.2 omicron reinfected, BA.2 convalescents, (Fig. S11 N, Q, J)) and individual landscapes of the two BA.2 convalescent samples with high titers against non-omicron variants (G778, G780). The slope of the BA.2 conv. landscape, excluding the two cross-reactive samples, was set to 1, all other slopes and serum coordinates were fitted as described in *Material & Methods.* The map shown in panel **B** serves as base plane, the z-axis reflects $log_2$ titers with each two-fold increase marked starting from titer 20. The grey plane at titer 50 serves as reference.

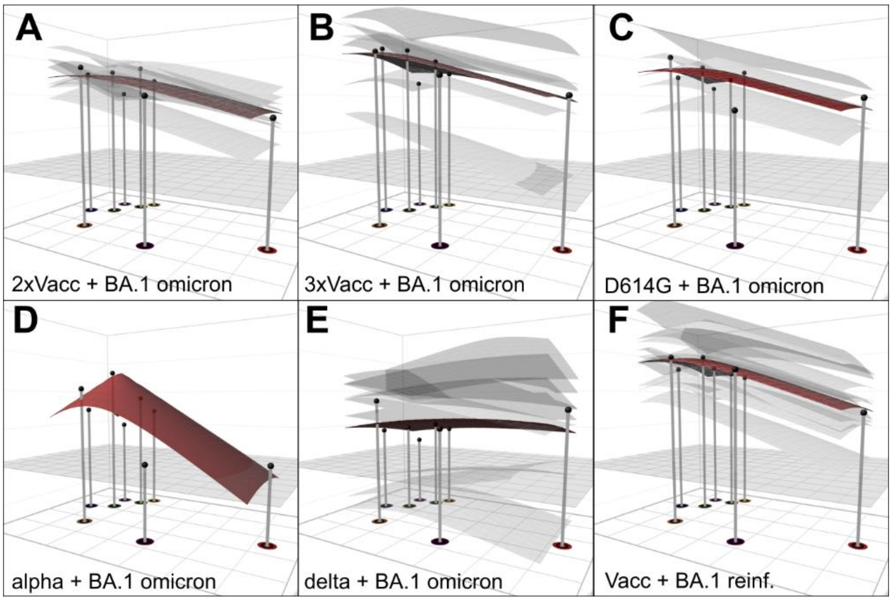

**Figure S13. Variability of BA.1 omicron multi-exposure landscapes.** Individual (transparent) and GMT (solid) antibody landscapes for the Vacc + BA.1 omicron (**A-B)**, BA.1 omicron reinf. (**C-E)** and Vacc + BA.1 omicron reinf. (**D**) after subsetting by number of vaccinations and prior infection (Fig. S11 M, O). The map in Figure 2 serves as base plane, the serum coordinates per serum and landscape slopes per additional serum group were fitted as described in *Materials & Methods.* Serum reactivity bias was adjusted for GMT calculation as described in [3]. The z-axis reflects $log_2$ titers with each two-fold increase marked starting from titer 20. The grey plane at titer 50 serves as reference.

### Supplementary Tables

#### Table S1. Patient Characteristics

| Study cohort | number of participants | vaccination -(+/-) | Variant of infection | | Mean age (years) ± SD | % female (N) |
|---|---|---|---|---|---|---|
| | | | first | second | | |
| **Single exposure** | | | | | | |
| First wave convalescent | 10 | - | ancestral | - | 51.6 ± 11.8 | 60.0 (6) |
| Alpha convalescent | 10 | - | alpha | - | 49.1 ± 14.3 | 80.0 (8) |
| Beta convalescent | 8 | - | beta | - | 79.8 ± 9.5 | 50.0 (4) |
| Delta convalescent | 7 | - | delta | - | 27.3 ± 10.3 | 42.9 (3) |
| BA.1 convalescent | 18 | - | BA.1 omicron | - | 47.0 ± 17.2 | 61.1 (11) |
| BA.2 convalescent | 12 | - | BA.2 omicron | - | 35.8 ± 14.9 | 50.0 (6) |
| **Double exposure** | | | | | | |
| BA.1 reinfected | 15 | - | ancestral (n=5)<br>alpha (n=1)<br>delta (n=9) | BA.1 omicron | 35.0 ± 16.9 | 72.7 (8) |
| BA.2 reinfected | 3 | - | ancestral (n=1)<br>delta (n=2) | BA.2 omicron | 34.7 ± 6.2 | 66.7 (2) |
| mRNA-1273/ mRNA-1273 | 10 | + | - | - | 35.4 ± 9.4 | 60.0 (6) |
| AZ/AZ | 10 | + | - | - | 37.3 ± 9.3 | 80.0 (8) |
| AZ/BNT | 10 | + | - | - | 32.8 ± 4.1 | 70.0 (7) |
| BNT/BNT | 11 | + | - | - | 36.1 ± 10.6 | 63.6 (7) |
| **≥ Triple exposure** | | | | | | |
| Vacc + BA.1 | 14 | + | omicron (BA.1) | - | 36.5 ± 16.8 | 57.1 (8) |
| Vacc + BA.2 | 8 | + | omicron (BA.2) | - | 32.8 ± 4.1 | 62.5 (5) |
| Vacc + BA.1 reinfected | 11 | + | ancestral (n=4)<br>alpha (n=1)<br>delta (n=6) | BA.1 omicron | 36.4 ± 16.8 | 54.5 (6) |
| AZ/AZ + delta | 6 | + | delta | - | 30.3 ± 4.6 | 100.0 (6) |
| BNT/BNT + delta | 22 | + | delta | - | 50.7 ± 14.4 | 59.1 (13) |

**Table S2. Statistics for neutralizing antibody titers against indicated variant compared to BA.2 omicron.**

| | Comparison of BA.2 titers against$ | | | | | | |
|---|---|---|---|---|---|---|---|
| **Study cohort** | **D614G** | **Alpha** | **Alpha + E484K** | **Beta** | **Gamma** | **Delta** | **BA.1 Omicron** |
| **Single exposure** | | | | | | | |
| BA.2 convalescent | ns | * | * | *** | * | ns | ns |
| First wave convalescent | ** | * | ns | ns | ** | * | ns |
| **Double exposure** | | | | | | | |
| AZ/AZ + delta | ns | ns | *** | ns | ns | ns | ns |
| BNT/BNT + delta | **** | **** | *** | ns | **** | *** | ns |

$ using non-parametric repeated measures ANOVA with Friedman`s test for multiple comparisons

**Table S3. Representativeness of study.**

| | |
|---|---|
| Disease, problem, or condition under investigation | BA.2 omicron differs immunologically from both BA.1 omicron and pre-omicron variants |
| Special considerations related to | |
| Sex | Female study participants were slightly overrepresented. Seroprevalence through infection in Austria is similar among males and females.[4] A study found equal titers of neutralizing antibodies between male and female participants.[5] |
| Age | Age was similar in all vaccinated groups; however, in unvaccinated convalescent there were differences. This might influence absolute titers of neutralizing antibodies; however, the ratio between the different variants should not be affected by this. |
| Ethnicity | Majority of population in Tyrol/Austria is white |
| Geography | Rates of vaccination and prior infection vary throughout the world. Austria has a vaccination rate of 73.1% (29.04.2022).[6] Seroprevalence in Tyrol has increased in a study among blood donors (healthy adults 18-70 years) from 3.4% in June 2020 to 82.7% in September 2021.[7] |
| Durability of cross-neutralizing antibodies | Some of the samples from convalescent individuals were collected shortly after infection (2-4 weeks) and therefore conclusions over longevity of these antibodies are limited. |
| Overall representativeness of this study | Small sample size and retrospective study design are major limitations of the current study. Further studies are needed to include newer omicron sub-variants into the antigenic maps such as BA.4, BA.5 and BA.2.12.1, which are currently rising in South Africa and the USA. |

**Supplementary References**

1. Wilks S. Racmacs: R Antigenic Cartography Macros. (https://acorg.github.io/Racmacs/index.html).
2. Fonville JM, Wilks SH, James SL, et al. Antibody landscapes after influenza virus infection or vaccination. Science 2014;346(6212):996-1000. DOI: doi:10.1126/science.1256427.
3. Wilks SH, Mühlemann B, Shen X, et al. Mapping SARS-CoV-2 antigenic relationships and serological responses. bioRxiv 2022:2022.01.28.477987. DOI: 10.1101/2022.01.28.477987.
4. Knabl L, Mitra T, Kimpel J, et al. High SARS-CoV-2 seroprevalence in children and adults in the Austrian ski resort of Ischgl. Commun Med (London) 2021;1(1):4. (In eng). DOI: 10.1038/s43856-021-00007-1.
5. Pichler D, Baumgartner M, Kimpel J, et al. Marked Increase in Avidity of SARS-CoV-2 Antibodies 7-8 Months After Infection Is Not Diminished in Old Age. The Journal of infectious diseases 2021;224(5):764-770. (In eng). DOI: 10.1093/infdis/jiab300.
6. Our World in Data. (https://ourworldindata.org/covid-vaccinations#what-share-of-the-population-has-completed-the-initial-vaccination-protocol).
7. Siller A, Seekircher L, Wachter GA, et al. Seroprevalence, waning, and correlates of anti-SARS-CoV-2 IgG antibodies in Tyrol, Austria: Large-scale study of 35,193 blood donors conducted between June 2020 and September 2021. medRxiv 2021:2021.12.27.21268456. DOI: 10.1101/2021.12.27.21268456.
